# Ethical Risks and Governance of Consumer Electroencephalography-Based Neurotechnology: Qualitative Interview Study With Expert Stakeholders

**DOI:** 10.64898/2026.11.09.26367587

**Authors:** Julie Moonga, Leili Soo

## Abstract

**Background:** Consumer electroencephalography (EEG)-based neurotechnologies are rapidly expanding across commercial digital health delivery networks and wellness markets. Positioned at the intersection of non-medical self-monitoring and clinical applications, these consumer devices introduce complex challenges for device governance and digital bioethics regarding product claims, data privacy, evidential substantiation, and user protection.

**Objective:** This qualitative study examined expert stakeholder perspectives on the ethical, regulatory, and data governance challenges of consumer EEG neurotechnology, focusing on the allocation of responsibility across product lifecycles.

**Methods:** We conducted semi-structured interviews with 19 expert stakeholders spanning research, clinical practice, engineering, industry, ethics, law, and regulatory policy across North America, Europe, and Latin America. Interview data were systematically analysed using reflexive thematic analysis to examine how governance challenges, evidence burdens, and responsibilities are conceptualized across the neurotechnology ecosystem.

**Results:** Five interrelated themes emerged: (1) Regulatory Category Boundaries and Oversight; (2) Evidential Credibility and Claims-Related Harm; (3) Neurodata Privacy, Security, and Inference; (4) Layered Governance and Explicit Accountability; and (5) Participation, Global Equity, and Contextual Responsiveness. Participants identified major friction in applying traditional medical oversight to nonmedical digital health uses, linked unverified claims to user vulnerability, and expressed concern over downstream neurodata processing. A connecting synthesis demonstrates that oversight responsibilities span multiple actors and development stages rather than resting on a single regulatory mechanism.

**Conclusions:** Effective governance of consumer EEG digital health devices requires explicit allocation of duties for claims substantiation, data practices, and user safety, backed by inclusive stakeholder participation and contextual responsiveness.

**Author Summary:** Consumer electroencephalography (EEG) devices are becoming widely available to the public for focus, relaxation, and wellness tracking. While these non-invasive brain-computer interfaces offer exciting opportunities for digital health, they also introduce complex risks regarding product marketing, data privacy, and user safety. Because these devices often sit outside formal medical device regulations, it is unclear who is responsible for verifying their claims or protecting the sensitive data they generate.

In this study, we interviewed 19 experts across technology development, neuroscience, law, and regulatory policy from North America, Europe, and Latin America. We explored how these stakeholders understand the risks of consumer neurotechnology and where responsibility should lie. Our findings show that governance cannot rely on a single law or voluntary industry standard alone. Instead, protecting users requires a layered approach where obligations for proof, data handling, and ongoing monitoring are explicitly assigned to companies, platforms, and regulators. Additionally, governance must include the voices of users and patients to ensure digital neurotechnologies are developed and used responsibly across diverse global settings.

## Introduction

### Background

Consumer neurotechnology has expanded from specialized research and clinical environments into commercial markets. EEG-based headsets and related non-invasive brain-computer interface applications are promoted for meditation, self-monitoring, cognitive enhancement, and entertainment [1,2]. Some products are positioned as wellness technologies, while others make claims that approach health-related assessment or intervention. Their ethical significance therefore depends on their purposes, claims, and uses, alongside their technical characteristics.

Consumer EEG systems can combine recording hardware with applications, computational models, and data-processing services. These systems may translate physiological signals into feedback that users interpret as information about attention, relaxation, or other aspects of their mental functioning [3]. Evaluating such products requires distinguishing the ability to record a signal from the evidence supporting a particular interpretation. Brain-based feedback may influence users’ expectations and self-understanding even when its reliability or practical significance is uncertain [1]. Accordingly, the relevant object of governance includes the hardware, software, product claims, data practices, and circumstances of use.

### Prior Work and Research Gap

Existing scholarship has identified ethical concerns involving evidential quality, privacy, security, autonomy, and oversight in direct-to-consumer neurotechnology [1,2]. Evidence and claims are closely connected: users may have limited means to assess whether advertised benefits or interpretations are adequately substantiated. These concerns are particularly consequential when products appeal to people seeking help with health-related difficulties. Ethical assessment must therefore consider both technical performance and how uncertainty is communicated.

Data governance presents related challenges. Concerns extend beyond the initial collection of EEG signals to their processing, linkage with other information, sharing, and subsequent use [3,4]. The significance of data can depend on the models applied to them and the purposes for which derived outputs are used. Research on BCI data governance has also highlighted questions about ownership, privacy, and responsibility [5]. These concerns do not imply that EEG provides unrestricted access to mental states. They concern the protection of information and the justification of interpretations within wider technological and commercial systems.

Several governance domains may address aspects of these products. Medical-device and general-wellness policies attend to intended purposes and claims, while data-protection and AI frameworks address aspects of processing and system use where applicable [6-8]. Technical standards and organizational processes offer additional approaches to ethical design and implementation [9]. However, the presence of multiple instruments leaves practical questions about how responsibilities connect across hardware, software, claims, and data pathways. Determining which rules apply is therefore only part of the governance problem.

Expert interviews offer a vital empirical tool for examining how these responsibilities are understood across the broader technology ecosystem. In digital health ecosystems, where hardware, cloud infrastructure, third-party applications, and user data streams intersect, governance cannot rely solely on static regulatory frameworks or proprietary closed-door practices. Instead, establishing effective oversight requires insights into how multi-sector actors conceptualize data privacy, open science principles, and practical technological implementation. Such accounts also require critical interpretation because professional roles inevitably shape judgments about risk and appropriate intervention. Responsible innovation scholarship further emphasizes the importance of including affected people in decisions about emerging technologies [10-12]. Expert perspectives can illuminate these governance questions but cannot substitute for users’ experiences. However, prior research has rarely triangulated perspectives across the full spectrum of digital health stakeholders. An unresolved question is how stakeholders working across the neurotechnology ecosystem—integrating insights across industry developers, clinical researchers, and policy experts spanning diverse geographical and regulatory environments—understand the allocation of responsibility when product claims, data practices, and oversight involve multiple organizations.

### Study Objective

This qualitative interview study investigated expert stakeholder perspectives on the ethical risks, data privacy challenges, and governance of consumer EEG-based neurotechnology. Drawing on semi-structured interviews with 19 cross-sector experts, including software and hardware developers, clinicians, researchers, bioethicists, and regulatory policy experts across North America, Europe, and Latin America, the study examined how stakeholders navigate tensions between open technology ecosystems and user protection, evaluating product claims, neurodata handling, and the allocation of governance responsibilities across the product lifecycle.

## Methods

### Study Design

We conducted a qualitative interview study to examine expert stakeholder perspectives on the ethical, legal, and regulatory challenges of consumer EEG-based neurotechnologies and non-invasive brain-computer interfaces. Semi-structured interviews systematically explored perceived risks, limitations of existing oversight, product claims, data privacy practices, and the allocation of governance responsibility.

### Participants and Recruitment

Nineteen expert stakeholders participated in the study. Participants were selected for their professional expertise or experience in neuroscience, clinical practice, neuroengineering, device development, data science, philosophy, ethics, law, regulation, or policy. Recruitment combined purposive and snowball sampling across North America, Europe, and Latin America. Potential participants were identified through searches of institutional databases and recommendations generated through snowball recruitment. They were invited by email, and interview dates and times were confirmed through email correspondence. Recruitment sought diversity in professional background, gender, and seniority. Recruitment ended after 19 interviews when the researchers judged that additional interviews were no longer contributing new concepts or relationships relevant to the study aims.

Participants were organized into three broad categories according to their primary area of expertise: researchers and clinicians, including philosophers and data-informatics specialists; engineers, developers, and industry stakeholders; and stakeholders working in ethics, law, regulation, and policy. These categories contextualized participants’ accounts and supported interpretation of how professional roles shaped their perspectives, functioning as qualitative context rather than statistically comparable groups.

### Data Collection

Individual semi-structured interviews were conducted in English using licensed Zoom or Microsoft Teams platforms, according to participant preference. Interviews lasted approximately 30 to 45 minutes. A common interview guide was used across stakeholder groups, allowing participants to address the questions from their own professional perspectives.

The guide covered three broad areas: participants’ professional background and work with neurotechnology; perceived benefits, risks, and barriers to application; and ethical, legal, and regulatory issues and potential governance responses. Topics included device safety, accuracy, efficacy, privacy, security, responsibility, policy gaps, standards, and mechanisms for preventing harm. Participants were also asked how ethical considerations informed their work and what barriers they encountered in practice.

Julie Moonga conducted all interviews. Interviews were audio-recorded, and transcripts were generated during the interviews using the videoconferencing platforms’ transcription functions. The transcripts formed the textual dataset for analysis.

### Data Analysis

Authors J.M. and L.S. analyzed the interview data using thematic analysis. Analysis followed an iterative process of familiarization, coding, theme development, review, and interpretation, drawing on the approach described by Braun and Clarke [13-16]. Analysis focused on patterns of meaning across participants’ accounts rather than fine-grained linguistic or conversational features.

Transcripts were read repeatedly to identify relevant passages and develop initial codes. Coding captured concerns about safety, accuracy, efficacy, misleading claims, privacy, security, vulnerability, responsibility, regulation, standards, transparency, and inequality. Participant responses were also condensed into progressively more abstract summaries to examine relationships among concerns and develop broader analytical categories. Codes and categories were compared across the dataset and revised as analysis progressed.

Related codes were grouped into candidate themes. These themes were reviewed against the coded extracts and the wider dataset, and their boundaries and definitions were refined to produce five final themes. Analysis examined both the concerns participants expressed and how they located responsibility for addressing them. Particular attention was given to relationships among product classification, evidential credibility, data practices, user protection, and governance mechanisms.

Coding discrepancies were reviewed to reach agreement on the interpretation of relevant extracts. Quotations were selected to illustrate recurring patterns, tensions, and analytically important positions. Minor edits removed repeated filler words or irrelevant fragments without changing meaning. Quotations are attributed using anonymized participant codes.

### Interpretation and Reflexivity

Participants’ accounts were interpreted in relation to their professional roles and responsibilities. The analysis considered how disciplinary and institutional perspectives could influence judgments about acceptable risk, adequate evidence, and appropriate oversight. Accounts were therefore treated as situated expert perspectives rather than definitive assessments of device performance, legal applicability, or consumer experience.

The interpretation was informed by neuroethics and governance scholarship, including attention to relationships among technologies, institutions, commercial actors, and users. Theme development remained responsive to the interview material, and candidate interpretations were reviewed against participants’ accounts. The policy implications presented in the Discussion constitute the authors’ synthesis of the findings in relation to the literature and are distinguished from the interview findings.

### Ethical Considerations

The study received ethical approval from King’s College London (United Kingdom) under reference LRS-22/23-39469. Participants provided verbal consent before interview recording. Participant identities are protected through anonymized quotation identifiers and broad reporting of professional categories. Full transcripts are not publicly available because of the sensitivity of the interviews and the restrictions associated with ethical approval. Deidentified quotations are presented to substantiate the analysis.

## Results

### Participant Characteristics

Nineteen expert stakeholders participated: 5 were categorized as researchers or clinicians, 8 as engineers, developers, or industry stakeholders, and 6 as stakeholders working in ethics, law, regulation, or policy. Participants worked across North America, Europe, and Latin America. Table 1 summarizes their professional backgrounds using broad categories to protect confidentiality.

**Table 1.** Participant characteristics and stakeholder categories. Stakeholders were grouped according to their primary area of expertise. Categories are intentionally broad to protect participant confidentiality.

| **Stakeholder category** | **Examples of expertise** | **Number of participants** |
|---|---|---|
| Researchers/clinicians | Neuroscience, neurology, psychiatry, psychology, clinical research, philosophy, data informatics | n = 5 |
| Engineers/developers/industry | Neuroengineering, device development, manufacturers | n = 8 |
| Ethics/law/regulation/policy | Neuroethics, law, regulatory policy, governance | n = 6 |
| Total |  | n = 19 |

### Overview of Themes

Thematic analysis generated five interrelated themes concerning regulatory boundaries, evidential credibility, neurodata governance, accountability, and participation. Across these themes, participants systematically evaluated consumer EEG applications, connecting technical device characteristics with product claims, potential uses, institutional responsibilities, and user protections.

A connecting interpretation was that oversight difficulties extended beyond identifying which rules applied. Participants also raised questions about who should substantiate claims, protect data, prevent misuse, and respond to users’ needs. Table 2 summarizes the themes and their central analytical interpretations.

**Table 2.** Themes, central analytical interpretations, and illustrative concerns identified through thematic analysis.

| **Theme** | **Central analytical interpretation** | **Illustrative concerns** |
|---|---|---|
| Theme 1: Regulatory Category Boundaries and Oversight | Participants perceived uncertainty about how oversight should follow neurotechnologies across clinical, wellness, enhancement, and other nonmedical contexts. | Medical and nonmedical boundaries; enhancement; breadth of applications; pace of technological change |
| Theme 2: Evidential Credibility and Claims-Related Harm | Accuracy and substantiation of claims were understood as matters of user protection, particularly when users sought health-related benefits. | Accuracy and precision; unsupported claims; changing brain states; vulnerability; limited information |
| Theme 3: Neurodata Privacy, Security, and Inference | Participants located data-governance concerns in the purposes, handling, interpretation, and potential reuse of data, as well as their collection. | Tracking; dual use; data value; purposes of collection; downstream use; model transparency |
| Theme 4: Layered Governance and Explicit Accountability | Participants proposed complementary governance mechanisms while questioning whether rules and standards alone could secure compliance and responsibility. | Soft law; statutory regulation; technical standards; professional guidance; company policies; enforcement |
| Theme 5: Participation, Global Equity, and Contextual Responsiveness | Participants associated meaningful governance with inclusion of affected people and attention to differences in cultural context and regulatory protection. | Patient participation; collaboration; sociocultural relevance; unequal protection; Global North–Global South differences |

### Theme 1: Regulatory Category Boundaries and Oversight

Participants described difficulties in applying established oversight approaches to technologies used across clinical and nonmedical settings. Their concerns involved the boundaries between treatment, wellness, and enhancement, alongside the growing range of applications. These boundaries were viewed as consequential for the protections available to users.

One participant contrasted established regulatory pathways with perceived uncertainty surrounding non-invasive technologies:

> "We must have rules … there is a regulatory system in place for that, unlike the non-invasive brain technology. Patient protection is my bigger concern in places where medicine is less regulated." (P1)

Another participant identified enhancement as a particular boundary problem:

> “We don’t have a regulatory system for enhancement.” (P2)

The pace and breadth of development also complicated participants’ understanding of oversight:

> "The technology is moving so fast and it is being used across our economy and society. It makes regulation hard, particularly the traditional way of regulating." (P3)

These accounts expressed perceived limitations of existing governance rather than establishing that non-invasive or enhancement technologies were legally unregulated. The analytical concern was how oversight could accommodate technologies whose applications extended beyond established clinical purposes. Participants’ accounts connected classification with the practical question of user protection: a familiar technology or an existing regulatory pathway did not, in their view, necessarily resolve the concerns arising from a different use.

### Theme 2: Evidential Credibility and Claims-Related Harm

Participants linked accuracy, precision, efficacy, and the substantiation of claims to protection of users. Technical performance was discussed in relation to what companies communicated and what users might expect from a device. Health-related claims were a particular concern because they could influence decisions by people seeking help.

One participant explicitly located responsibility for substantiating claims with companies:

> "The onus should be on the companies that are making claims. “Cure your depression”, for example, is one that I see all the time. If you are making that strong claim, it needs to be vetted." (P3)

The same participant emphasized that performance needed attention over time:

> "We need to get the right accuracy and precision, and then retrain over time to adjust to changing brain states." (P3)

A further account connected vulnerability with limited information:

> "When someone is a patient, desperation and lack of information can lead to decisions that are not so good." (P4)

Together, these extracts connected evidential quality with the circumstances in which users evaluated claims. Beyond whether a technology produced a raw output, the key analytical question was whether that output and its advertised meaning warranted users’ confidence. Participants’ concerns suggested that the burden of evaluating uncertain claims could fall on people who lacked sufficient information or were strongly motivated to obtain a benefit.

This theme therefore linked technical credibility with interpretive risk: the possibility that users would place confidence in claims or outputs whose evidential basis they could not readily assess. The interviews documented concerns regarding these potential risks, though quantifying the actual incidence of user harm remains beyond the study scope.

### Theme 3: Neurodata Privacy, Security, and Inference

Participants raised concerns about the collection, handling, interpretation, and potential reuse of brain-derived information. Their accounts connected privacy with tracking, dual use, the value of data, and the purposes for which information was processed.

One participant linked tracking with uncertainty about permissible uses:

> "Everything is tracking. What are the rules that are going to govern these sorts of things? Because we also have to deal with dual use." (P5)

Another emphasized the importance of examining purposes and subsequent use:

> “Policymakers and regulators should not only focus on data types but also why those data are collected and for what purposes, and how they will be used.” (P6)

Concerns also extended to the interpretation of data through computational models:

> “There’s no transparency into how the model after passing through all these hidden layers actually came to a particular conclusion.” (P3)

The latter account concerned model interpretation more broadly and illustrated a related problem for systems that translated data into user-facing conclusions. Participants’ concerns extended beyond initial data collection to encompass downstream processing, algorithmic conclusion generation, and secondary data uses.

The analytical interpretation was that data protection and interpretive accountability were connected. Decisions about collection, processing, and reuse could affect both privacy and the meaning attributed to an output. Rather than validating the empirical accuracy of consumer EEG in revealing sensitive mental states, these accounts focused on the governance of data and interpretations whose scope could expand beyond initial recordings.

### Theme 4: Layered Governance and Explicit Accountability

Participants described combinations of governance mechanisms rather than a single regulatory solution. Their proposals included standards, professional guidance, company policies, and statutory regulation. The value of combining mechanisms was accompanied by concern about implementation and compliance.

One participant described the need for complementary instruments:

> "We have no choice but to fill the gaps using soft law and maybe more than one mechanism, maybe standards, professional guidelines, company policies. It has to be some combination of those." (P7)

Another identified technical standards as an opportunity:

> "Opportunity is through standards. IEEE is looking at standards in the neuroscience or neural tech area, so that would be one opportunity to have an official industry standard, like an IEEE or ISO standard of appropriate governance." (P8)

However, a further participant challenged the assumption that legally binding rules necessarily resolved compliance problems:

> "Non-compliance is a problem not just with soft law, but also with hard law. There is tremendous non-compliance with hard law, so it is not unique to soft law." (P2)

These accounts revealed a tension between expanding the available governance instruments and ensuring that those instruments influenced practice. Standards and guidance were viewed as potential means of addressing gaps, while the account of noncompliance qualified any simple distinction between effective statutory regulation and ineffective voluntary approaches.

Read alongside the concerns about company claims and downstream data use, this theme raised the question of how responsibilities were allocated across governance arrangements. The analytical interpretation was that while multiple governance instruments may cover distinct technical domains, they risk obscuring underlying accountability unless explicit roles are defined. The interviews supported concern about the relationship between governance coverage, implementation, and accountability; they did not demonstrate that a particular combination of mechanisms would resolve that concern.

### Theme 5: Participation, Global Equity, and Contextual Responsiveness

Participants connected governance with the inclusion of affected people and attention to the contexts in which technologies were used. Their accounts questioned whether professional and technical expertise alone could adequately address patients’ perspectives and differences in regulatory protection.

One participant called for collaboration that included patients:

> "There must be a collaborative effort, not only with the clinicians, the researchers, developers and manufacturers, also with the patient, to really understand their culture and their humanistic perspective." (P1)

Another connected the value of neural data with unequal regulatory protection:

> "There are more regulations in the Global North than there are in the Global South, and neural data has a lot of value." (P3)

In a related extract, the same participant described differences in the perceived capacity to prevent misuse:

> "Global North are probably more aware of and can protect themselves in a way because there are regulations that are national regulations that can help prevent misuse of data." (P3)

These accounts linked participation and equity through a common concern about whose circumstances informed governance. Patient involvement was presented as a way to understand cultural and human perspectives, while the accounts of regulatory inequality drew attention to differences in available protection.

The analytical interpretation was that governance arrangements could not be assessed solely through their technical requirements or formal existence. Participants also considered whether affected people were included and whether protections were meaningful across settings. These were expert perspectives on participation and inequality, rather than direct evidence of consumer experience or a systematic comparison of jurisdictions.

## Discussion

### Principal Findings

This study examined expert stakeholder perspectives on the ethical risks and governance of consumer EEG-based neurotechnology. Five interrelated themes connected regulatory boundaries, evidential credibility, neurodata practices, accountability, and participation. Across these themes, participants considered both the availability of governance mechanisms and the responsibilities associated with their implementation.

The central interpretation is that consumer neurotechnology governance extends far beyond determining whether a device fits a traditional medical regulatory category. Because consumer EEG devices function within open and interconnected digital technology ecosystems, governance must address the full lifecycle—from hardware engineering and algorithmic model design to third-party data flows and end-user marketing. A key strength of this study lies in its multi-regional, cross-sector empirical design. By triangulating perspectives across 19 expert stakeholders spanning industry developers, clinical researchers, and policy authorities across North America, Europe, and Latin America, our findings reflect the real-world friction between open digital innovation and user safety. This cross-sector alignment reveals that layering voluntary guidelines, technical standards, and statutory rules is ineffective unless specific responsibilities for data privacy, model validation, and user protection are explicitly assigned to identifiable actors.

This interpretation should be distinguished from a demonstrated accountability failure. The study examined expert accounts rather than product compliance, enforcement decisions, or consumers’ experiences of harm. It nevertheless identifies questions that existing governance arrangements need to address: who substantiates a claim, who remains responsible for downstream data uses, and who responds when product functionality or interpretation changes? Fig 1 brings together the five themes and illustrates how concerns about evidence, data practices, and oversight connect with responsibility across the product lifecycle.

### Comparison With Prior Work

Previous scholarship has identified concerns about safety, privacy, autonomy, transparency, and evidential quality in direct-to-consumer neurotechnology [1,2]. The present findings connect these concerns through stakeholders’ accounts of governance and responsibility. Participants did not discuss evidence, data protection, and regulation solely as separate problems. Their accounts linked the credibility of claims with user vulnerability, the value of data with potential misuse, and the availability of standards with questions of compliance.

The theme of regulatory boundaries illustrates this relationship. Participants perceived difficulties when technologies moved between clinical, wellness, enhancement, and other nonmedical applications. These accounts should not be interpreted as evidence that consumer neurotechnology is categorically unregulated.

Rather, they suggest that the boundaries used to allocate oversight may not resolve all concerns associated with actual use. This is consistent with arguments that governance of direct-to-consumer neurotechnology requires attention to product claims and applications, alongside technical characteristics [2].

The findings also reinforce the ethical significance of evidential credibility. Participants located responsibility for substantiating claims with companies and connected limited information with users’ vulnerability. This extends the discussion of safety beyond immediate physical effects to the confidence users place in product claims and feedback. Kreitmair [1] similarly identifies epistemic considerations as an important dimension of consumer neurotechnology ethics. In this study, the relevant concern was whether users could reasonably evaluate the meaning and reliability of information presented as brain-based. The interviews support attention to this interpretive burden, although they do not establish the frequency or consequences of misleading feedback.

The neurodata theme likewise connects privacy with the purposes and interpretation of information. Previous work has examined privacy and security risks in BCI ecosystems and the challenges associated with neural-data ownership and use [3-5,17]. Participants’ accounts emphasized collection purposes, subsequent uses, and transparency of computational interpretation. Their concerns therefore support examining the data pathway rather than treating the initial recording as the sole object of governance. This interpretation is consistent with Chiappone et al [18], whose comparative analysis of US and EU consumer data protections argues that governance should attend to the purposes and consequences of data use rather than focusing exclusively on whether information is classified as ‘neural data’. This does not imply that EEG provides unrestricted access to mental states. The ethical question concerns the handling of information and the claims made about its significance, including when interpretations are uncertain.

Finally, participants’ calls for patient involvement and attention to unequal regulatory protection align with scholarship on public participation and responsible innovation [10-12]. Their accounts suggest that governance depends partly on whose perspectives inform definitions of benefit, acceptable risk, and adequate protection.

Because the sample comprised experts, these findings establish the importance participants attributed to inclusion; they do not substitute for research with consumers or affected communities.

### Implications for Policy and Practice

The findings support an approach that connects governance requirements with identifiable responsibilities. The following implications are the authors’ interpretation of the interview findings in relation to the literature. First, assessment should consider the relationship between product purpose, claims, functionality, and foreseeable use. A distinction between medical and nonmedical products remains important, but it should not be treated as a complete assessment of ethical significance. Manufacturers should clearly describe intended purposes and the limits of their claims, while relevant authorities should clarify how existing requirements apply. The findings do not justify applying identical requirements to all consumer EEG products. They support examining which obligations are appropriate to the consequences of particular claims and uses.

Second, claims substantiation and communication of uncertainty deserve explicit attention. Developers and manufacturers are better positioned than consumers to provide evidence about the performance and limitations of their products. Disclosure should distinguish what has been evaluated from what remains uncertain and should identify the populations and conditions to which the evidence applies. Stronger claims, especially those directed toward people seeking health-related benefits, warrant closer scrutiny. This proposal follows participants’ concerns about company responsibility and vulnerability; determining specific evidential thresholds would require further technical and regulatory work.

Third, data governance must embrace an ecosystem-wide view that balances data privacy with transparent, open science practices. In consumer digital health, responsibilities for data handling are distributed across hardware manufacturers, app platforms, cloud analytics providers, and third-party software developers. Safeguarding user trust in open technology ecosystems requires clear documentation of data provenance, model transparency, and restrictions on secondary data monetization. Policy interventions must mandate explicit data pathways and organizational accountability rather than burdening end-users with deciphering complex consent agreements for derived neurodata outputs.

Fourth, standards and professional guidance should be connected with implementation and accountability. Participants identified these mechanisms as useful complements to statutory regulation, while also noting that noncompliance occurs under both voluntary and binding arrangements. The existence of a standard is therefore not, by itself, evidence of adequate protection. Its value depends on what it requires, how adherence is assessed, and what follows when requirements are not met. Process-based approaches to ethical design and organizational governance may help make these responsibilities operational, but their effectiveness must be evaluated in practice [9,19].

These responsibilities also have a temporal dimension. Changes to software, processing, or product claims may alter what users receive after initial deployment. A lifecycle perspective therefore offers a practical extension of the findings: organizations should consider whether modifications require renewed evaluation, revised communication, or changes to safeguards. The interviews do not establish a specific monitoring regime, but participants’ concerns about changing performance and compliance provide a rationale for examining continuing responsibility.

Finally, participation should contribute to identifying consequences and priorities that expert assessment may overlook. Users, patients, and affected communities can inform the interpretation of benefits, burdens, and acceptable data practices. This involvement should extend beyond consultation that has no influence on decisions. Cross-jurisdictional collaboration should also attend to differences in regulatory capacity and available protection, without assuming that frameworks developed in one setting will transfer unchanged to another [20]. The study identifies these considerations as governance priorities; their practical implementation requires work with the affected groups themselves. Table 3 consolidates these proposed responses, identifies the actors involved, and outlines considerations for implementation.

**Table 3.** Author-proposed governance responses informed by the interview findings.

| **Interview finding** | **Proposed response** | **Actors involved** | **Implementation consideration** |
|---|---|---|---|
| Uncertainty about oversight across medical and nonmedical applications | Clarify applicable obligations in relation to intended purpose, claims, functionality, and foreseeable use | Manufacturers, relevant regulators, consumer-protection authorities | Requirements should be proportionate to risk and assessed under applicable jurisdictional rules |
| Concern about unsupported claims, accuracy, and user vulnerability | Substantiate claims and communicate the scope and limitations of supporting evidence | Developers, manufacturers, distributors, relevant oversight authorities | Evidential scrutiny should reflect the strength and consequences of claims |
| Concern about collection purposes, downstream use, and interpretation | Document data pathways, assign responsibilities, and scrutinize subsequent uses and derived outputs | Manufacturers, platforms, developers, organizations processing data | Safeguards should address organizational practices as well as user consent |
| Support for complementary instruments alongside concern about compliance | Connect standards and guidance with assessment of adherence and mechanisms for responding to failures | Standards bodies, manufacturers, professional organizations, oversight authorities | Adoption of an instrument should not be equated with effective implementation |
| Calls for patient participation and attention to unequal protection | Involve affected groups in governance decisions and assess applicability across contexts | Developers, researchers, regulators, users, patients, community representatives | Participation should influence decisions; transfer across settings requires local assessment |

### Limitations

While our sample (N = 19) provided deep qualitative triangulation across three distinct stakeholder sectors (developers/industry, clinicians/researchers, and ethics/policy experts) across three major global regions (North America, Europe, and Latin America), it does not represent all regional regulatory environments or consumer end-user populations systematically. The multidisciplinary breadth supported robust cross-sector policy insights, but dedicated comparative studies across specific legal jurisdictions remain necessary.

The analysis examined participants’ accounts rather than independently assessing device performance, legal compliance, enforcement, or harm. Proposed governance responses consequently remain implications for further consideration, rather than demonstrated solutions. In addition, product functionality and governance arrangements can change over time. Applying these findings to particular technologies or jurisdictions requires attention to the circumstances in which the interviews were conducted and subsequent developments.

Future research should examine consumer experiences, specific product and data pathways, and how responsibilities are allocated in practice. Evaluations of standards, claims review, and mechanisms for responding to concerns would help establish whether the proposed responses improve protection.

## Conclusions

Expert stakeholders connected the governance of consumer EEG-based neurotechnology with regulatory boundaries, evidential credibility, neurodata practices, accountability, and participation. Their accounts suggest that ethical assessment should follow the relationships among products, claims, data uses, interpretations, and users. Combining governance mechanisms may address different concerns, but their effectiveness depends on clear responsibilities and implementation. The practical implication is to make responsibility for evidence, data practices, and user protection explicit, while involving affected people in determining what adequate protection requires.

## Data Availability

Data available but stored under UK GDPR

## Acknowledgements

We thank the interview participants for their time and for sharing their perspectives and expertise.

## Figure Captions

**Fig 1. Synthesis of the five interview themes, the connecting interpretation, and the authors’ proposed governance implication.** Connecting lines indicate analytical relationships rather than causal effects.

## Notes

### Competing Interest Statement

No conflict of interest

### Author Declarations

The study received ethical approval from King's College London (United Kingdom) under reference LRS-22/23-175 39469.

## References

1. Kreitmair KV. Dimensions of ethical direct-to-consumer neurotechnologies. AJOB Neurosci. 2019;10(4):152–166. doi:10.1080/21507740.2019.1665120. PMID:31642755.

2. Wexler A, Reiner PB. Oversight of direct-to-consumer neurotechnologies. Science. 2019;363(6424):234–235. doi:10.1126/science.aav0223. PMID:30655433.

3. Ienca M, Haselager P, Emanuel EJ. Brain leaks and consumer neurotechnology. Nat Biotechnol. 2018;36(9):805-810. doi:10.1038/nbt.4240. PMID:30188521.

4. Bonaci T, Calo R, Chizeck HJ. App stores for the brain: privacy and security in brain-computer interfaces. IEEE Technol Soc Mag. 2015;34(2):32–39. doi:10.1109/MTS.2015.2425551.

5. Naufel S, Klein E. Brain-computer interface (BCI) researcher perspectives on neural data ownership and privacy. J Neural Eng. 2020;17(1):016039. doi:10.1088/1741-2552/ab5b7f. PMID:31766026.

6. US Food and Drug Administration. General wellness: policy for low risk devices. Guidance for industry and Food and Drug Administration staff. January 6, 2026. URL: https://www.fda.gov/regulatory-information/search-fda-guidance-documents/general-wellness-policy-low-risk-devices [accessed 2026-10-01].

7. European Parliament, Council of the European Union. Regulation (EU) 2016/679 of the European Parliament and of the Council of 27 April 2016 on the protection of natural persons with regard to the processing of personal data and on the free movement of such data, and repealing Directive 95/46/EC (General Data Protection Regulation). Off J Eur Union. 2016;L119:1–88. URL: https://eur-lex.europa.eu/eli/reg/2016/679/oj/eng [accessed 2026-10-01].

8. European Parliament, Council of the European Union. Regulation (EU) 2024/1689 of the European Parliament and of the Council of 13 June 2024 laying down harmonised rules on artificial intelligence and amending Regulations (EC) No 300/2008, (EU) No 167/2013, (EU) No 168/2013, (EU) 2018/858, (EU) 2018/1139 and (EU) 2019/2144 and Directives 2014/90/EU, (EU) 2016/797 and (EU) 2020/1828 (Artificial Intelligence Act). Off J Eur Union. 2024;L2024/1689. URL: https://eur-lex.europa.eu/eli/reg/2024/1689/oj/eng [accessed 2026-10-01].

9. IEEE Standards Association. IEEE standard model process for addressing ethical concerns during system design. IEEE Std 7000-2021. IEEE; 2021. doi:10.1109/IEEESTD.2021.9536679.

10. Jasanoff S. Technologies of humility: citizen participation in governing science. Minerva. 2003;41(3):223–244. doi:10.1023/A:1025557512320.

11. Owen R, Macnaghten P, Stilgoe J. Responsible research and innovation: from science in society to science for society, with society. Sci Public Policy. 2012;39(6):751–760. doi:10.1093/scipol/scs093.

12. Stilgoe J, Owen R, Macnaghten P. Developing a framework for responsible innovation. Res Policy. 2013;42(9):1568–1580. doi:10.1016/j.respol.2013.05.008.

13. Braun V, Clarke V. Using thematic analysis in psychology. Qual Res Psychol. 2006;3(2):77–101. doi:10.1191/1478088706qp063oa.

14. Braun V, Clarke V. Thematic analysis. In: Cooper H, Camic PM, Long DL, Panter AT, Rindskopf D, Sher KJ, editors. APA Handbook of Research Methods in Psychology. Vol 2: Research Designs: Quantitative, Qualitative, Neuropsychological, and Biological. Washington, DC: American Psychological Association; 2012:57–71. doi:10.1037/13620-004.

15. Braun V, Clarke V. Reflecting on reflexive thematic analysis. Qual Res Sport Exerc Health. 2019;11(4):589–597. doi:10.1080/2159676X.2019.1628806.

16. Braun V, Clarke V. Thematic Analysis: A Practical Guide. London, UK: SAGE Publications; 2022. ISBN:9781473953246.

17. Ienca M, Haselager P. Hacking the brain: brain-computer interfacing technology and the ethics of neurosecurity. Ethics Inf Technol. 2016;18(2):117–129. doi:10.1007/s10676-016-9398-9.

18. Chiappone N, Urian D, Soo L, Matney C, Moonga J, Rosero-Villarreal F, et al. The illusion of distinction in ‘neural data’ governance: rethinking United States consumer data protections through a European lens. Neuroethics. 2026;19:36. doi:10.1007/s12152-026-09659-z.

19. Coglianese C, Lazer D. Management-based regulation: prescribing private management to achieve public goals. Law Soc Rev. 2003;37(4):691–730. doi:10.1046/j.0023-9216.2003.03703001.x.

20. Organisation for Economic Co-operation and Development. Recommendation of the Council on Responsible Innovation in Neurotechnology. OECD/LEGAL/0457. 2019. URL: https://legalinstruments.oecd.org/en/instruments/OECD-LEGAL-0457 [accessed 2026-10-01].

